# The Impact of the Temperature on Extracting Information From Clinical Trial Publications Using Large Language Models

**DOI:** 10.1101/2024.10.23.24316005

**Authors:** Paul Windisch, Fabio Dennstädt, Carole Koechli, Christina Schröder, Daniel M. Aebersold, Robert Förster, Daniel R. Zwahlen

## Abstract

**Introduction:** The application of natural language processing (NLP) for extracting data from biomedical research has gained momentum with the advent of large language models (LLMs). However, the effect of different LLM parameters, such as temperature settings, on biomedical text mining remains underexplored and a consensus on what settings can be considered “safe” is missing. This study evaluates the impact of temperature settings on LLM performance for a named-entity recognition and a classification task in clinical trial publications.

**Methods:** Two datasets were analyzed using GPT-4o and GPT-4o-mini models at nine different temperature settings (0.00–2.00). The models were used to extract the number of randomized participants and classified abstracts as randomized controlled trials (RCTs) and/or as oncology-related. Different performance metrics were calculated for each temperature setting and task.

**Results:** Both models provided correctly formatted predictions for more than 98.7% of abstracts across temperatures from 0.00 to 1.50. While the number of correctly formatted predictions started to decrease afterwards with the most notable drop between temperatures 1.75 and 2.00, the other performance metrics remained largely stable.

**Conclusion:** Temperature settings at or below 1.50 yielded consistent performance across text mining tasks, with performance declines at higher settings. These findings are aligned with research on different temperature settings for other tasks, suggesting stable performance within a controlled temperature range across various NLP applications.

## Introduction

Using natural language processing (NLP) to extract data from biomedical research publications has seen increasing interest following the development of more powerful architectures, most notably large language models (LLMs).^1–3^ The task itself has been of interest for a long time as the ability to automatically extract and structure information, e.g. according to PICO (patient, intervention, control, outcome) characteristics, could improve various processes such as screening the literature for relevant publications, assessing adherence to reporting guidelines and ultimately automating the process of evidence synthesis.^4,5^

While the capability of LLMs for several of these text mining tasks has been demonstrated previously, there is still relatively little information on the impact that different parameters or prompts might have.^6,7^ A notable parameter of LLMs is the temperature of its softmax function. A high temperature leads to a flat probability distribution for the prediction of the next token which makes the model more likely to choose less conventional options.^8^ This may increase the creativity of the output but also makes the behavior less predictive while increasing the risk of incoherent output. With a low temperature setting, the model will choose only the most likely next token thus leading to a more predictive, coherent output with limited creativity. While anecdotal reports of decreasing performance for certain tasks at higher temperatures exist, there are also publications reporting consistent performance across a broad temperature range e.g. for answering multiple choice questions or predicting clinical outcomes like in-hospital mortality from electronic health records.^9,10^ Thus, there is no consensus on what temperature range can be considered “safe”, i.e. which range is unlikely to result in decreasing performance.

The purpose of this project was therefore to evaluate the impact of the temperature setting on text mining tasks for clinical trial publications to assess if there is also a constant performance across a wider range of temperatures as has been demonstrated for other tasks and beyond which threshold the performance starts to drop. As most text mining tasks generally fall into the categories of either named-entity recognition or classification, we used two dedicated datasets for these tasks.

## Methods

Two datasets that had been annotated as part of previous projects by the author group were used to create tasks for the evaluation of two LLMs, namely Generative Pretrained Transformer 4 Omni (GPT-4o, OpenAI, San Francisco, United States) and GPT-4o mini at nine different temperature settings (0.00, 0.25, 0.50, 0.75, 1.00, 1.25, 1.50, 1.75, 2.00).^11–14^ The respective versions that were used were gpt-4o-2024-05-13 and gpt-4o-mini-2024-07-18.

The first task was to extract the number of people who underwent randomization from the abstract of a publication reporting on a randomized clinical trial (RCT). To this end, a random sample of 996 randomized controlled trials (RCTs) from seven major journals (British Medical Journal, JAMA, JAMA Oncology, Journal of Clinical Oncology, Lancet, Lancet Oncology, New England Journal of Medicine) published between 2010 and 2022 were labeled. The abstracts were retrieved as a txt file from PubMed and parsed using regular expressions (i.e., expressions that match certain patterns in text). For each trial, the number of randomized trial participants was retrieved by looking at the abstract, followed by the full publication if the number could not be determined with certainty from the abstract. Two physician annotators carried out the annotation independently and conflicts were resolved by discussing the differences afterwards.

The LLMs were called via the application programming interface (API) with the aforementioned temperatures and max_tokens set to 10 to stop the LLM in case of hallucinations. All other API-parameters were left at their default. The system prompt was the following: “*You will be provided with the abstract of a randomized controlled clinical trial. Your task will be to extract the number of people who underwent randomization. If this number is not explicitly mentioned, you may use other numerical information (e*.*g. the number of total participants or adding up the number of patients in each arm). Please return only the number as a single integer. If no information is available, please return null*.”

The user prompt was the respective abstract. The raw responses were stored and afterwards, each raw response was converted into an integer unless the conversion failed, e.g. due to the raw response being equal to “null” or due to non-numerical hallucinations.

The results were evaluated against the ground truth created by the human annotators. The percentage of correctly formatted, numerical predictions was calculated as well as performance metrics like the mean absolute percentage error (MAPE) and the proportion of predictions that fell within a certain percentage of the ground truth.

The second task was to classify an abstract regarding whether or not it reported on an RCT and/or an oncology topic. To this end, a random sample of 900 publications from the aforementioned seven major journals published between 2010 and 2022 were annotated. Publications that described RCTs received the label “RCT”. Publications that covered oncological topics received the label “ONCOLOGY”. Trials that fulfilled both criteria were assigned both labels. Trials that were neither RCTs nor covered oncology topics were assigned no label. The two labels were chosen as each label poses different requirements to the LLM: For the oncology label, the model does not need a deep contextual understanding but can rather make a prediction based on the presence of certain words that are associated with oncology publications, such as “cancer” or words related to staging and antineoplastic therapies. In order to assign the RCT label, the model can not simply rely on the presence of words and phrases like “randomized” or “primary endpoint” as these might also be present in other articles such as meta-analyses of randomized controlled trials.

Annotation was based on the title and abstract, which were also retrieved as a txt file from PubMed and parsed using regular expressions. Due to the relatively simple annotation process, annotation was carried out by a single physician annotator. The API call to the LLMs used the same settings as for the first task. The user prompt was again the abstract. The system prompt was the following: “*You will be provided with the abstract of a medical publication. Your task will be to determine if the abstract reports on a randomized controlled trial. If the abstract reports on a systematic review or meta-analysis of randomized controlled trials or a commentary/editorial, return false. In addition, you will be asked to determine if the abstract focusses on an oncology topic which includes all papers dealing with the prevention, diagnosis or treatment of solid or hematologic cancers. Your response should be a list of two boolean values (True or False), the first indicating if the paper is an RCT and the second indicating if the paper is oncology-related. The list should be enclosed in brackets and separated by a comma, e*.*g. [True, False]*.”

The raw responses were stored and afterwards, the two boolean values were extracted unless the extraction failed due to incorrect formatting. The results were evaluated against the ground truth by computing the proportion of correctly formatted predictions as well as the confusion matrices for each label and several performance metrics (accuracy, precision, recall, and F1 score).

All programming was performed in Python (version 3.11.5) using, among others, the pandas (version 2.1.0) and openai (version 1.40.3) packages.

## Results

The median number of people who underwent randomization was 668 with an interquartile range (IQR of 300 - 1836) and a histogram of the respective number of people who underwent randomization in each trial is presented in Figure 1A. The percentage of trials with correctly formatted numerical predictions made by GPT-4o was almost constant between temperatures 0.00 and 1.50, ranging from 98.7% to 99.0%. The first noticeable drop occurred at temperature 1.75 with 95.6% with a further drop to 89.2% at temperature 2.00. The same pattern could be seen with GPT-4o mini where temperatures between 0.00 and 1.50 resulted in trials with correctly formatted numerical predictions between 99.0% and 99.1% and drops at 1.75 as well as 2.00 (97.5% and 90.2% respectively). A scatterplot of the predictions of the LLMs compared to the ground truth is presented in Figure 1B. The complete performance metrics are presented in Table 1.

**Table 1.**
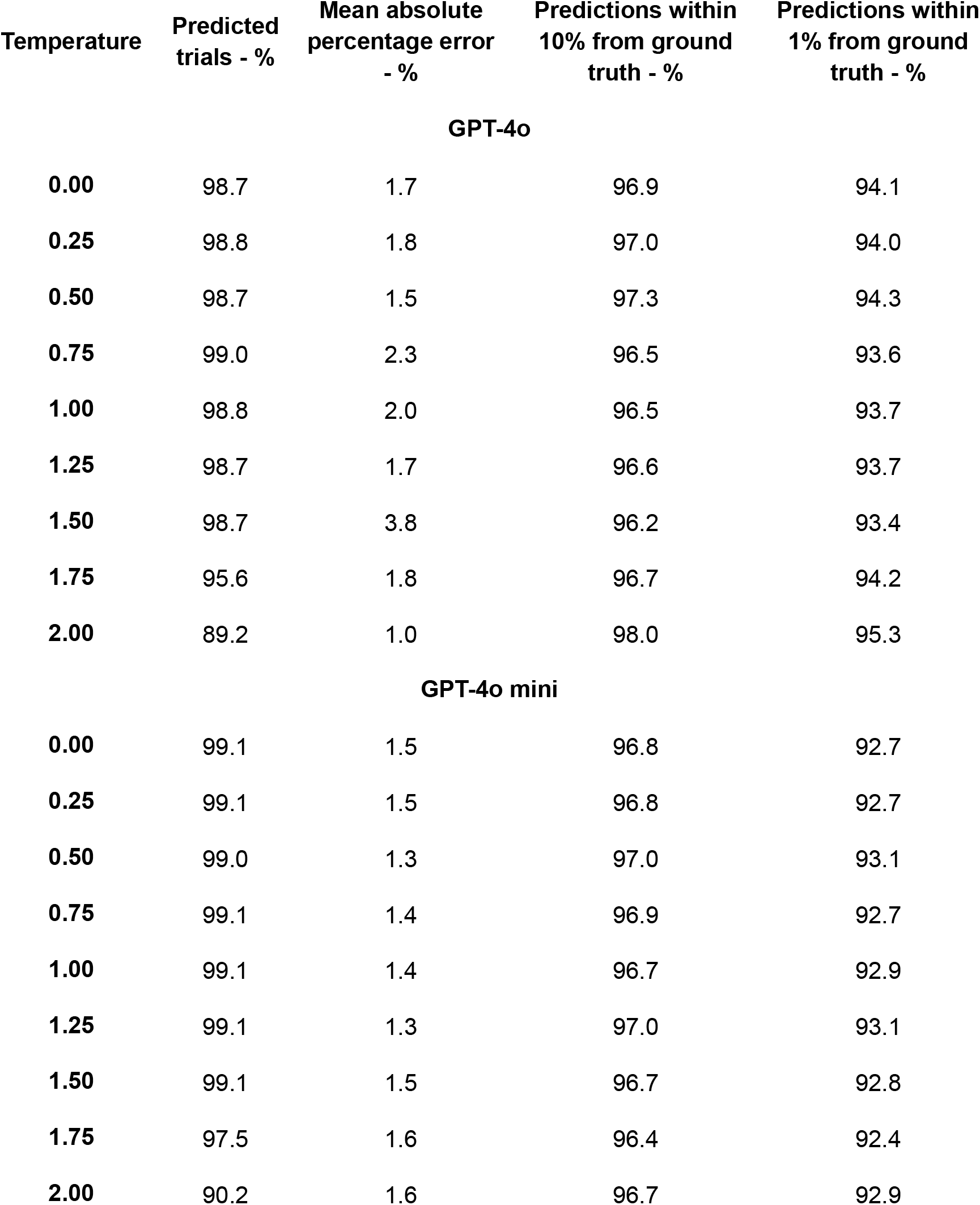
Performance of GPT-4o and GPT-4o mini when asked to extract the number of patients who were randomized from the abstract of a publication reporting on a randomized controlled trial. “Predicted trials” indicates the percentage of trials for which a correctly formatted, numerical prediction was returned.

| Temperature | Predicted trials - % | Mean absolute percentage error - % | Predictions within 10% from ground truth - % | Predictions within 1% from ground truth - % |
| --- | --- | --- | --- | --- |
| **GPT-4o** |  |  |  |  |
| **0.00** | 98.7 | 1.7 | 96.9 | 94.1 |
| **0.25** | 98.8 | 1.8 | 97.0 | 94.0 |
| **0.50** | 98.7 | 1.5 | 97.3 | 94.3 |
| **0.75** | 99.0 | 2.3 | 96.5 | 93.6 |
| **1.00** | 98.8 | 2.0 | 96.5 | 93.7 |
| **1.25** | 98.7 | 1.7 | 96.6 | 93.7 |
| **1.50** | 98.7 | 3.8 | 96.2 | 93.4 |
| **1.75** | 95.6 | 1.8 | 96.7 | 94.2 |
| **2.00** | 89.2 | 1.0 | 98.0 | 95.3 |
| **GPT-4o mini** |  |  |  |  |
| **0.00** | 99.1 | 1.5 | 96.8 | 92.7 |
| **0.25** | 99.1 | 1.5 | 96.8 | 92.7 |
| **0.50** | 99.0 | 1.3 | 97.0 | 93.1 |
| **0.75** | 99.1 | 1.4 | 96.9 | 92.7 |
| **1.00** | 99.1 | 1.4 | 96.7 | 92.9 |
| **1.25** | 99.1 | 1.3 | 97.0 | 93.1 |
| **1.50** | 99.1 | 1.5 | 96.7 | 92.8 |
| **1.75** | 97.5 | 1.6 | 96.4 | 92.4 |
| **2.00** | 90.2 | 1.6 | 96.7 | 92.9 |

**Figure 1.**
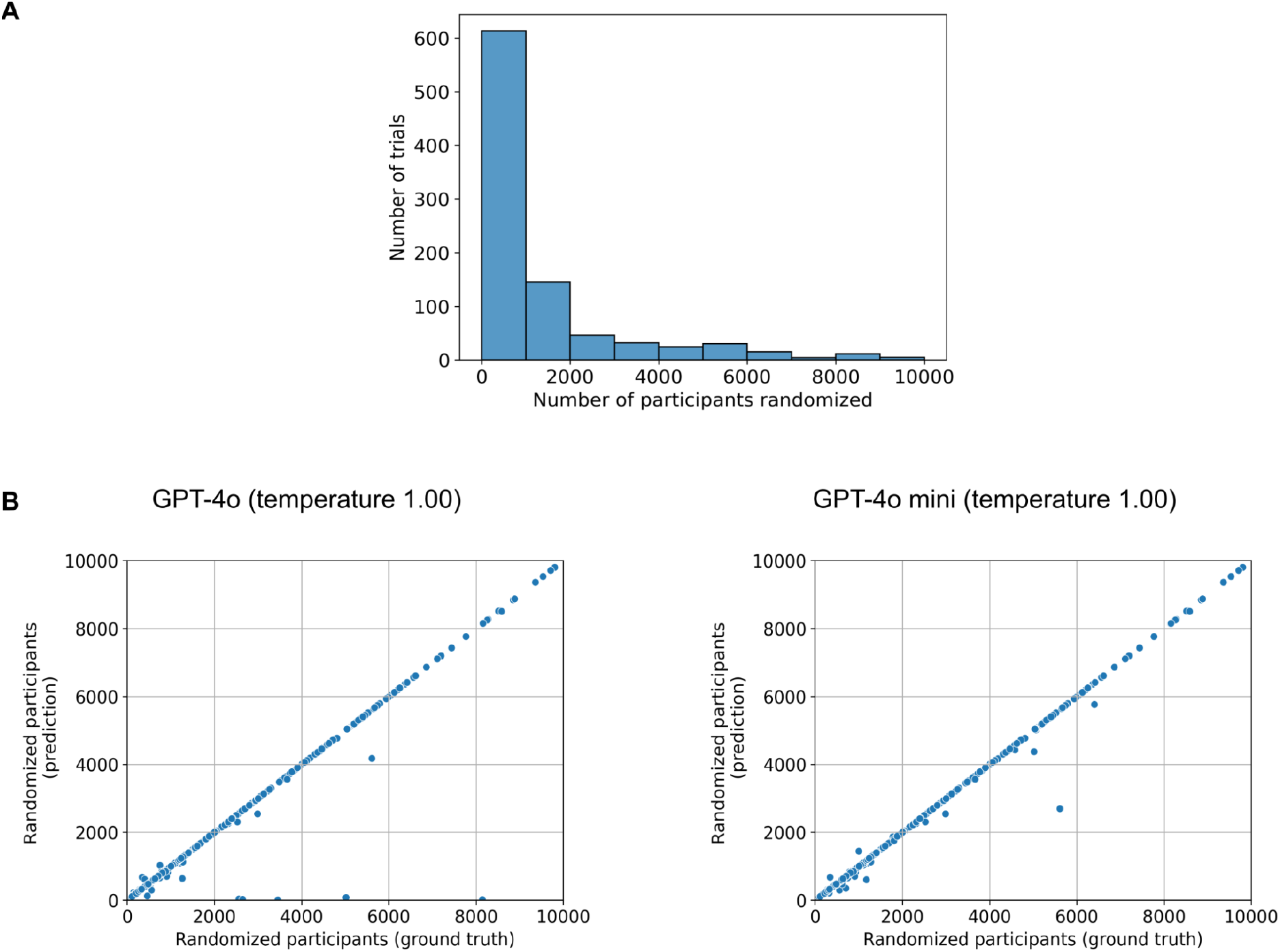
Predicting or extracting the number of randomized trial participants. A) Histogram of the trials and the respective number of randomized participants. B) Scatterplots where each dot represents a trial with its x-coordinate representing how many people were randomized and the y-coordinate representing what GPT-4o (left) or GPT-4o mini (right) predicted in terms of how many people were randomized at temperature 1.00. To ensure better visualization of the range that most trials fall into, only trials and predictions with less than 10’000 randomized participants are displayed.

When analyzing only the correctly formatted predictions, the performance in terms of mean absolute percentage errors (MAPE) and the proportion of predictions within a certain margin of error did not show a major drop beyond a certain temperature. On the contrary, for GPT-4o, a temperature of 2.00 resulted in the lowest MAPE and the highest proportion of predictions within 10% and 1% of the ground truth.

A confusion matrix on the distribution of RCTs and oncology trials is presented in Figure 2A. 46.8% of trials were RCTs and 26.9% covered an oncology topic. The predictions of the LLMs compared to the ground truth for each label are presented in the confusion matrices in Figure 2B. The performance metrics are presented in Tables 2 (RCT) and 3 (oncology). The classification task resulted in trials with correctly formatted predictions in 100% of abstracts with both models and labels for temperatures at or below 1.25. The biggest drop occurred again between temperatures 1.75 (98.9% for GPT-4o and 97.7% for GPT-4o mini) and 2.00 (93.3% for GPT-4o and 92.4% for GPT-4o mini) for both labels. The F1 scores for the label RCT ranged from 0.956 to 0.960 for GPT-4o and 0.914 to 0.921 for GPT-4o mini. The F1 scores for the label oncology ranged from 0.965 to 0.977 for GPT-4o and 0.964 to 0.972 for GPT-4o mini.

**Table 2.** Performance of GPT-4o and GPT-4o mini when asked to predict if an abstract of a publication reported on a randomized controlled trial. “Predicted trials” indicates the percentage of trials for which a correctly formatted prediction was returned.

| Temperature | Predicted trials - % | Accuracy | Precision | Recall | F1 Score |
| --- | --- | --- | --- | --- | --- |
| **GPT-4o** |  |  |  |  |  |
| **0.00** | 100.0 | 0.957 | 0.915 | 1.000 | 0.956 |
| **0.25** | 100.0 | 0.959 | 0.919 | 1.000 | 0.958 |
| **0.50** | 100.0 | 0.957 | 0.915 | 1.000 | 0.956 |
| **0.75** | 100.0 | 0.958 | 0.917 | 1.000 | 0.957 |
| **1.00** | 100.0 | 0.960 | 0.921 | 1.000 | 0.959 |
| **1.25** | 100.0 | 0.960 | 0.921 | 1.000 | 0.959 |
| **1.50** | 99.9 | 0.961 | 0.923 | 1.000 | 0.960 |
| **1.75** | 98.9 | 0.960 | 0.921 | 1.000 | 0.959 |
| **2.00** | 93.3 | 0.957 | 0.916 | 1.000 | 0.956 |
| **GPT-4o mini** |  |  |  |  |  |
| **0.00** | 100.0 | 0.915 | 0.847 | 1.000 | 0.917 |
| **0.25** | 100.0 | 0.918 | 0.851 | 1.000 | 0.919 |
| **0.50** | 100.0 | 0.915 | 0.847 | 1.000 | 0.917 |
| **0.75** | 100.0 | 0.919 | 0.852 | 1.000 | 0.920 |
| **1.00** | 100.0 | 0.920 | 0.854 | 1.000 | 0.921 |
| **1.25** | 100.0 | 0.917 | 0.849 | 1.000 | 0.918 |
| **1.50** | 99.6 | 0.918 | 0.852 | 1.000 | 0.920 |
| **1.75** | 97.7 | 0.916 | 0.849 | 1.000 | 0.918 |
| **2.00** | 92.4 | 0.911 | 0.842 | 1.000 | 0.914 |

**Table 3.** Performance of GPT-4o and GPT-4o mini when asked to predict if an abstract of a publication reported on an oncology topic. “Predicted trials” indicates the percentage of trials for which a correctly formatted prediction was returned.

| Temperature | Predicted trials - % | Accuracy | Precision | Recall | F1 Score |
| --- | --- | --- | --- | --- | --- |
| **GPT-4o** |  |  |  |  |  |
| **0.00** | 100.0 | 0.983 | 0.967 | 0.971 | 0.969 |
| **0.25** | 100.0 | 0.982 | 0.963 | 0.971 | 0.967 |
| **0.50** | 100.0 | 0.984 | 0.967 | 0.975 | 0.971 |
| **0.75** | 100.0 | 0.983 | 0.963 | 0.975 | 0.969 |
| **1.00** | 100.0 | 0.982 | 0.963 | 0.971 | 0.967 |
| **1.25** | 100.0 | 0.983 | 0.963 | 0.975 | 0.969 |
| **1.50** | 99.9 | 0.981 | 0.963 | 0.967 | 0.965 |
| **1.75** | 98.9 | 0.988 | 0.967 | 0.988 | 0.977 |
| **2.00** | 93.3 | 0.985 | 0.970 | 0.974 | 0.972 |
| **GPT-4o mini** |  |  |  |  |  |
| **0.00** | 100.0 | 0.983 | 0.952 | 0.988 | 0.970 |
| **0.25** | 100.0 | 0.983 | 0.952 | 0.988 | 0.970 |
| **0.50** | 100.0 | 0.984 | 0.956 | 0.988 | 0.972 |
| **0.75** | 100.0 | 0.982 | 0.945 | 0.992 | 0.968 |
| **1.00** | 100.0 | 0.984 | 0.956 | 0.988 | 0.972 |
| **1.25** | 100.0 | 0.984 | 0.956 | 0.988 | 0.972 |
| **1.50** | 99.6 | 0.980 | 0.937 | 0.992 | 0.964 |
| **1.75** | 97.7 | 0.981 | 0.940 | 0.992 | 0.965 |
| **2.00** | 92.4 | 0.983 | 0.953 | 0.987 | 0.970 |

**Figure 2.**
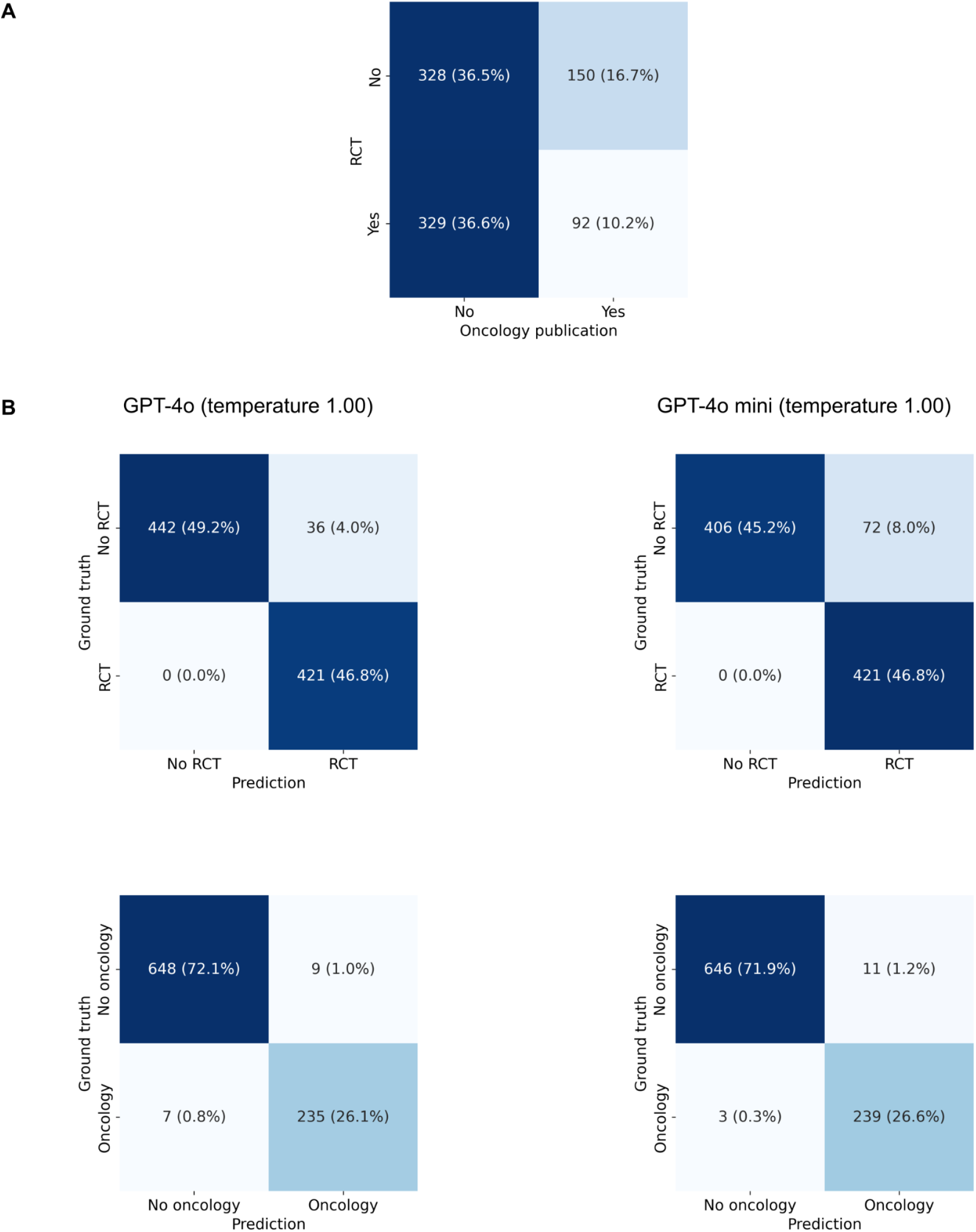
Trial classification. A) Confusion matrix regarding whether or not the abstract of a publication reports on a randomized controlled trial and whether or not it reports on an oncology topic. B) Confusion matrices for the predictions of GPT-4o (left) or GPT-4o mini (right) at a temperature of 1.00 regarding whether or not the abstract of a publication reports on a randomized controlled trial (top) and whether or not it reports on an oncology topic (bottom).

## Discussion

In this study, temperatures at or below 1.50 yielded comparable results and the most pronounced drop in performance occurred between temperatures 1.75 and 2.00. Notably, the drop in performance only occurred with regard to the proportion of correctly formatted predictions while the error metrics of the correctly formatted predictions remained constant and in some cases even improved. A possible explanation for this seemingly counterintuitive improvement could be that abstracts where the number of people randomized is not explicitly stated and the model has to try to infer it from the information it has, are morelikely to trigger a hallucination. Thus, more difficult abstracts end up with incorrectly formatted predictions which results in better performance when looking only at the correctly formatted predictions. These findings regarding text mining are largely consistent with Patel and colleagues who saw a consistent performance of various LLMs (GPT-4, GPT-3.5, and Llama-3-70b) for various clinical task across a temperature range from 0.2 to 1.0 as well as Renze and colleagues who saw consistent results for an even wider array of LLMs between temperatures of 0.0 to 1.0.^9,10^

A second insight of this observation is that specifying a desired output format via the prompt could facilitate the detection of hallucinations. This then raises the question if forcing the LLM to adhere to a certain output format e.g. using features such as Structured Outputs or JSON mode could lead to this behavior being removed and more incorrect predictions ending up being correctly formatted. This topic warrants further research.

Another interesting observation is that there was a difference in performance between GPT-4o and GPT4o-mini for predicting whether or not an abstract reports on a randomized controlled trial (RCT), but a similar performance for predicting whether the abstract covers an oncology topic. A possible explanation for this is that determining if an abstract reports on an oncology topic is the simpler task as it mainly requires “knowing” which words in an abstract are connected to oncology. To determine if an abstract reports on an RCT, one cannot only rely on the terminology but has to understand the context. As an example, the phrase “randomized controlled trial” can occur in an abstract reporting on a randomized controlled trial, but also in a systematic review that included randomized controlled trials. Therefore, it seems plausible that the more powerful model, i.e. GPT-4o, performs better at this task.

This study is limited by the fact that only OpenAI models were used to analyze the different temperature settings. However, the evaluation studies of different temperatures settings for other tasks such as those mentioned previously indicate that the findings are likely to generalize to other architectures.^9,10^ Furthermore, other factors influencing the output of the LLM, such as the choice of model, additional model parameters or different prompts, were not investigated. The specific setting of these factors can influence each other as well as the output and, in turn, the performance. While it seems unlikely that changes in these factors would yield substantially different results, one should be aware of this fact when interpreting this study and likely do at least a focused evaluation of different parameters for the task that one is trying to accomplish.

As a potential outlook, one could try to create more robust text mining workflows by sending the same or slightly varied prompts to models with different temperature settings in the seemingly safe range of temperatures from 0.00 to 1.50. If all models are in agreement, the prediction is considered correct. If there is a disagreement, manual review is triggered. This workflow could also be implemented to reduce costs if, for example, three predictions from small, cheaper models at different temperatures are requested and a more expensive model is only used if there is a disagreement.

In conclusion, temperature settings at or below 1.50 seem to result in comparable performance for extracting information from medical publications. These findings are aligned with research on different temperature settings for other tasks, suggesting that a safe temperature range may be consistent across a variety of applications.

## Data Availability

All data and code used for the analysis have been uploaded to https://github.com/windisch-paul/temperature. The dataset has also been submitted to Dryad and is currently undergoing review.

https://github.com/windisch-paul/temperature

## Notes

**Competing interests:** P.W. has a patent application titled ‘Method for detection of neurological abnormalities’ outside of the submitted work. The remaining authors declare no conflict of interest.

### Competing Interest Statement

P.W. has a patent application titled 'Method for detection of neurological abnormalities' outside of the submitted work. The remaining authors declare no conflict of interest.

### Clinical Protocols

https://github.com/windisch-paul/temperature

### Funding Statement

This study did not receive any funding

